# Intraluminal Prucalopride Increases Propulsive Motor Activities in the Human Colon

**DOI:** 10.1101/2020.05.24.20111930

**Authors:** Mitra Shokrollahi, Xuan-Yu Wang, Natalija Milkova, Jan D Huizinga, Ji-Hong Chen

## Abstract

**Background:** Luminal application of 5-HT_4_ receptor agonists can increase peristalsis in the guinea pig, mouse, rat and rabbit colon. Our aim in the present study was to test the effects of intraluminal prucalopride on motor patterns in the human colon.

**Methods:** Colonic motor patterns were studied in vivo in a healthy volunteer using High-Resolution Colonic Manometry (HRCM) with an 84-sensor water perfused catheter with 1cm spacing. 5-HT and 5-HT_4_ receptor immunohistochemistry was performed on human tissue biopsies throughout the colon.

**Key results:** Activating mucosal 5-HT_4_ receptors via intraluminal prucalopride enhanced propulsive motor activity in the human colon by increasing occurrence and amplitude of propulsive motor patterns including high-amplitude propagating pressure waves (HAPWs), pancolonic simultaneous pressure waves (SPWs) and HAPW-SPWs. Prucalopride-induced motor patterns had a close temporal association with a significant degree of anal sphincter relaxation and some were accompanied by a strong urge to defecate. Biopsies showed 100% colocalization of the 5-HT_4_ receptor to enterochromaffin cells throughout the colon and rectum.

**Conclusions and inferences:** Activating luminal 5-HT_4_ receptors on enterochromaffin cells by intraluminal prucalopride increased propulsive motor activity. 5-HT_4_ receptors were found only on enterochromaffin cells and not ubiquitous on all epithelial cells. Our data support incorporation of prucalopride in colon-specific drug delivery systems as a prokinetic to treat colonic hypomotility disorders.

**50 word abstract:** High-resolution colonic manometry and biopsy immunohistochemistry revealed that 5-HT_4_ receptors in the lumen of the human colon are present exclusively on enterochromaffin cells and that the 5-HT_4_ agonist prucalopride evokes all major propulsive motor patterns, associated with significant anal sphincter relaxation, when given intraluminally.

**250-character clinical message:** Activating luminal 5-HT_4_ receptors on enterochromaffin cells by intraluminal prucalopride increased propulsive motor activity in the human colon. Colon-specific delivery systems with a 5-HT_4_ agonist may become the preferred colon prokinetic.

## INTRODUCTION

5-HT_4_ receptors (5-HT_4_R) have long been among the most compelling targets for prokinetic agents. Oral doses of 5-HT_4_R agonists have been used clinically to treat dysmotility; clinical trials have provided evidence that the 5-HT_4_R agonist prucalopride, given orally, increases transit in humans and can be effective in managing constipation (3, 5, 8). Corroborating live animal studies have shown that oral or intravenous prucalopride result in enhancement of peristaltic activity in the dog colon (6). Studies on whole rat colon by our team provided further insights into actions of 5-HT on colonic motor patterns (51, 52). Furthermore, using high-resolution colonic manometry (HRCM), we observed that oral intake of prucalopride can initiate peristaltic activity within 10 minutes in human healthy volunteers (37) likely through a gastro-colonic reflex mediated by gastric 5-HT_4_Rs. However, despite the clinical success of 5-HT_4_R agonists, concerns about their adverse cardiovascular side effects and inconsistent efficacy has limited their use. Several mechanisms of action may underly 5-HT_4_R agonists because of the fact that within the GI tract, 5-HT_4_Rs are widespread among various types of enteric neurons, 5-HT containing enterochromaffin cells, smooth muscle cells and interstitial cells of Cajal (ICC). Moreover, 5-HT_4_R expression is different between anatomical regions and among different species (22).

Animal models have demonstrated that luminal application of 5-HT_4_R agonists promotes propulsive motility in the gastrointestinal tract (26) (49) (19) (36). For instance, mucosal application of the 5-HT_4_R partial agonist tegaserod promoted peristalsis in rat and guinea pig colonic segments (21). Moreover, pellet propulsion studies showed that administration of intraluminal 5-HT_4_R agonists tegaserod, prucalopride and naronapride increased the velocity of pellet propulsion significantly more than their serosal application in guinea pig distal colon. This suggests that intraluminal administration of 5-HT_4_R agonists may be more effective than their oral application in regulating propulsive activity (26) (36) (21) (27). Mucosal application of tegaserod activated 5-HT release from enterochromaffin cells, mucus discharge from goblet cells and Cl^−^ secretion from enterocytes in mouse and guinea pig colonic tissue segments as well as human biopsies (26). Such responses were generated in a TTX insensitive and antagonist sensitive manner indicating that 5-HT_4_R agonists stimulated 5-HT release from within the lumen by directly activating EC cells rather than via a neural mechanism (26) (45) (22).

Altered 5-HT signaling has been associated with chronic constipation and irritable bowel syndrome (IBS). El-Salhy and colleagues reported that the number of 5-HT-immunoreactive cells per unit area of epithelial cells is lower in colons of patients with slow transit constipation compared to healthy controls (17), suggesting a role for 5-HT signaling in gut homeostasis (2) (47, 48) (32). Consistent with animal studies mentioned above, we have recently shown that intraluminal perfusion of the highly selective 5-HT_4_R agonist prucalopride significantly increases propulsive motor activities in the whole proximal and mid rabbit colon (45). This likely occurs via 5-HT release from enterochromaffin cells into the lamina propria to activate intestinal primary afferent neurons (IPANs) and subsequently the myenteric motor neurons (22).

Propulsive activity in the rabbit colon takes the form of the Colonic Motor Complexes (CMCs) (15, 24, 33, 39, 45). The development of the CMC occurs in a dose-dependent and antagonist sensitive manner, manifested in various levels of excitation beginning with clusters of fast propagating contractions followed by long distance contractions (LDCs) as the most forceful representation of the CMC in the rabbit. Additionally, intraluminal prucalopride and intraluminal exogenous 5-HT significantly increased contraction amplitude, intraluminal pressure amplitude, frequency, velocity and degree of propagation of the CMC along the colon (45).

In this case study, we show that intraluminal prucalopride enhances propulsive motor activities in the human colon.

## MATERIALS AND METHODS

### Study Subject

A 24-year-old female healthy volunteer was recruited through local advertising. The participant gave written informed consent and all procedures were approved by the Hamilton Integrated Research Ethics Board (HiREB). Exclusion criteria were abdominal surgery, hepatic, kidney or cardiac diseases, connective tissue disorders, central nervous system disorders, thyroid diseases, prostate diseases or malignancies. The subject had normal stool consistency and normal bowel frequency, between 1 every 3 days and 3 per day. The subject had no defecation difficulty and was not taking any medication.

### High- Resolution Colonic Manometry (HRCM)

High-resolution colonic manometry was performed on a custom-made platform (Medical Measurement Systems (MMS); Laborie, Toronto, ON, Canada). An 84-sensor water-perfused 105 cm long catheter was designed (Mui Scientific, Mississauga, ON, Canada). The spacing between sensor 1-48 was 1.5 cm while the spacing between the rest of the sensors was 1 cm. The catheter included one 10-cm long balloon between sensors 7 and 8. Sensor 1 was placed in the ascending colon. The catheter was inserted with minimal sedation (fentanyl i.v. 50 mcg and midazolam i.v. 2 mg) with the assistance of a colonoscope after a bowel cleaning procedure using an inert osmotic laxative (PEG-Lyte, Pendopharm, Montreal, QC, Canada), but no use of stimulant laxatives such as bisacodyl. For the bowel cleaning procedure, 3L of PEG (70 g/L) was taken between 4 and 6 pm the day before the procedure, with more water consumed as needed to have all solids removed. The next morning, 1L was taken at 4 am. The tip of the catheter was clipped to the mucosa via a fish line, a few centimeters distal to the cecum. The catheter was made of 100% silicon. After use, an extensive approved cleaning procedure was executed followed by sterilization. The subject was in the supine position during the entire recording, except during meal intake. Subject was instructed to report all events such as gas or liquid expulsion, bowel movements, pain, and discomfort. Subject was asked not to promote or prevent gas or liquid expulsion by increasing abdominal pressure or contracting the external anal sphincter if an urge arose. Artifacts caused by body movements such as changing body position, talking, coughing, laughing, and urination were noted immediately into the data acquisition files and excluded from the analysis.

#### Protocol

A 90-minute recording of baseline activity was started 30 minutes after the colonoscope was withdrawn. Afterwards, the response to a 5 min balloon distention at the proximal colon was evaluated. A 1000 kcal meal (500 g of organic vanilla yogurt fortified with organic milk fat (Mapleton Organics, Moorefield, ON, Canada) was given and its effect was observed for 90 min. Thereafter the effects of 2mg of prucalopride (Resotran) given in the proximal colon via the catheter were observed for 60 minutes. The prucalopride suspension was made in saline by crushing a tablet with a pestle and mortar for 5 minutes and suspending in 20 ml sterile water. Drug administration was followed by more water to flush the drug delivery catheter. At the end of the study, an X-ray was taken using a portable X-ray machine that was brought into the study room. The catheter which was used in this patient contains radiopaque markers which are visible under X-ray and were used to visualize the placement of the catheter along the colon.

#### Water-Perfusion

The catheter delivered 0.1 ml/min sterile water via each sensor to a total of 0.57 L per hour. A drainage tube (OD 3.3 mm) was placed in the rectum to allow passive outflow of excess water. The remainder of the water was expelled by colon motor activities or absorbed. Intraluminal pressure in between motor patterns did not change during the recording session (38), hence, the water inflow did not cause passive tonic pressure changes.

#### Quantification and Statistical analysis of HRCM Data

All data were stored using the software developed by Medical Measurement Systems (MMS; Laborie, Toronto, ON, Canada), and analyzed using programs developed by Sean Parsons by Image J (National Institutes of Health, Bethesda, MD, United States) and MATLAB (Mathworks, Natick, MA, United States). After HAPWs were identified visually in Image J, they were encircled free hand and a Contourer plug-in was used to set a 20 mmHg isobar and accurately outline the HAPW to obtain its amplitude as an average of all measured points within the contour. This method is substantially different from measurements using low-resolution manometry where only a few points along the motor pattern are considered. Other motor patterns such as SPWs were measured through freehand outline using a rectangular selection tool to obtain different characteristics such as amplitude, duration, propagation length and velocity, if applicable. Responses to stimuli were described and compared to baseline activity and activity after meal. Data are given as mean ± SEM. Significance was determined by ANOVA with multiple comparisons as mentioned in the table footnotes using Prism 8 software (GraphPad, United States), P < 0.05 was considered significant. Percentage of anal sphincter relaxation was calculated by comparing the mean amplitude of the anal sphincter in a 3-minute period before relaxation and the whole period of relaxation. Due to the oscillatory nature of the anal sphincter pressure, relaxation was quantified when it reached higher than 25% of the average pressure value recorded in the 3-minute period before the relaxation. The period of relaxation started with the first sign of relaxation and ended with return to its baseline pressure. In the middle of this period, complete anal sphincter relaxation could be achieved. MATLAB was used to generate the 3D images.

#### Identified Motor Patterns by HRCM

The focus of this case study was on propulsive motor patterns and their coordination with anal sphincter relaxation. Therefore, the following motor patterns were studied:

1. Simultaneous pressure waves (SPWs) are pressure transients that occur simultaneously at all sensors that record them, as identified by Rao et al., (41) and De Schryver et al., (14), and further defined and characterized by us (9, 10, 38). They have also been called simultaneous contractions (40) or pan-colonic pressurizations (11).
2. High-amplitude propagating pressure waves (HAPWs) are defined as transient increases in pressure of more than 50 mmHg that propagate almost always in anal direction (37). They are also called high-amplitude propagating contractions or sequences (4).
3. High-amplitude propagating pressure waves followed by simultaneous pressure waves (HAPW-SPWs). A proximal HAPW can promptly switch to an SPW at the transverse or descending colon (9) (37). The SPW within this motor pattern is not pan-colonic.
4. The colo-anal reflex: anal sphincter activity and its relaxation associated with the motor patterns described above (37).

#### Immunohistochemistry for 5-HT_4_R and 5-HT Double Labeling

Biopsies were taken in the proximal, transverse, descending colon and rectum and were fixed in 4% paraformaldehyde in 0.1M phosphate buffer (pH 7.4) for 24 hours at 4°C and processed for paraffin embedding. Paraffin sections of 4*μ*m were cut, mounted on coated slides, de-waxed and re-hydrated. Antigen retrieval was performed on paraffin sections prior to immunostaining by heating the slides in 0.01 M citrate buffer (pH 6.0) for 20 minutes. Non-specific binding was blocked in 5% normal goat serum (NGS, Sigma, St. Louis, MO) for one hour at room temperature. Sections were then incubated in primary antibodies overnight at room temperature and secondary antibodies for one hour at room temperature. Primary antibodies were polyclonal rabbit anti-5-HT_4_ receptor (Atlas Antibodies, Cat # HPA040591, Lot # R36821, 1:200) and monoclonal mouse anti-5-HT (ThermoFisher Scientific, Cat # MA5-12111, 1:100); secondary antibodies were Alexa 488 goat anti-rabbit IgG (Jackson ImmunoResearch, 1:300) for 5-HT4R staining and Cy3 goat anti-mouse IgG (Jackson ImmunoResearch, 1:200) for 5-HT staining. After rinsing several times, sections were mounted with fluoroshield mounting medium with DAPI (Abcam, Cat # ab104139) for nuclear counterstain. All antibodies were diluted in 0.05M PBS (pH 7.4) with 0.03% triton-X-100. Negative controls included the omission of primary antibodies from the incubation solution.

#### Immunohistochemistry for Rabbit anti 5-HT and Mouse anti 5-HT Double Labeling

The same paraffin sections and the same staining protocol as mentioned above were applied for 5-HT double labelling to investigate 100% colocalization of two 5-HT markers. Primary antibodies were rabbit anti human 5-HT (ImmunoStar, Cat # 20080, Lot # 1431001, 1:2000) and mouse anti-5-HT (ThermoFisher Scientific, Cat # MA5-12111, 1:100); secondary antibodies were Alexa 488 goat anti-rabbit IgG (Jackson ImmunoResearch, 1:300) for rabbit 5-HT staining and Cy3 conjugated goat anti-mouse IgG (Jackson ImmunoResearch, 1:200) for mouse 5-HT staining.

#### Immunohistochemistry for mouse anti 5-HT with Protease XXV for Enzyme-Induced Epitope Retrieval and Heat-Induced Epitope Retrieval with citrate buffer

Adjacent thin sections of 2 *μ*m were studied for mouse anti 5-HT staining with two different antigen retrieval techniques. Enzyme-Induced epitope retrieval with protease XXV was recommended by the manufacturer but we also performed heat-induced epitope retrieval to be able to double label with anti 5-HT_4_R. We added this test to observe whether the two techniques produced the same staining results.

All immunostaining was examined with a Leica DMRXA2 microscope with x20 objective lens. All pictures were taken using a Retiga Imaging digital camera attached to the microscope and an Apple computer with Volocity software (Improvision Inc., Montreal, QC). For each colon biopsy section, six to eight areas covering the whole section were selected.

## RESULTS

### Colonic Activities Before Administration of Luminal Prucalopride

HAPWs were not present at baseline nor postprandially while SPWs were observed at baseline (n=1) and after meal intake (n=3) (Figure 1A,B). At baseline, a pan-colonic SPW with a mean amplitude of 8.7 mmHg, maximum pressure of 24.0 mmHg and a duration of 3.7 s was observed, not associated with gas or liquid expulsion. After the meal, pan-colonic SPWs were observed with an amplitude of 7.2 ± 0.8 mmHg and duration of 12.2 ± 2.5 s which entered the rectum but were not associated with gas nor liquid expulsion (Table 1). The SPW observed at baseline was associated with 69% relaxation of the anal sphincter while SPWs evoked postprandially were associated with 87% relaxation. The anal sphincter showed an intrinsic rhythmicity of 5.5 cpm (Figures 1A,B).

**Figure 1.**
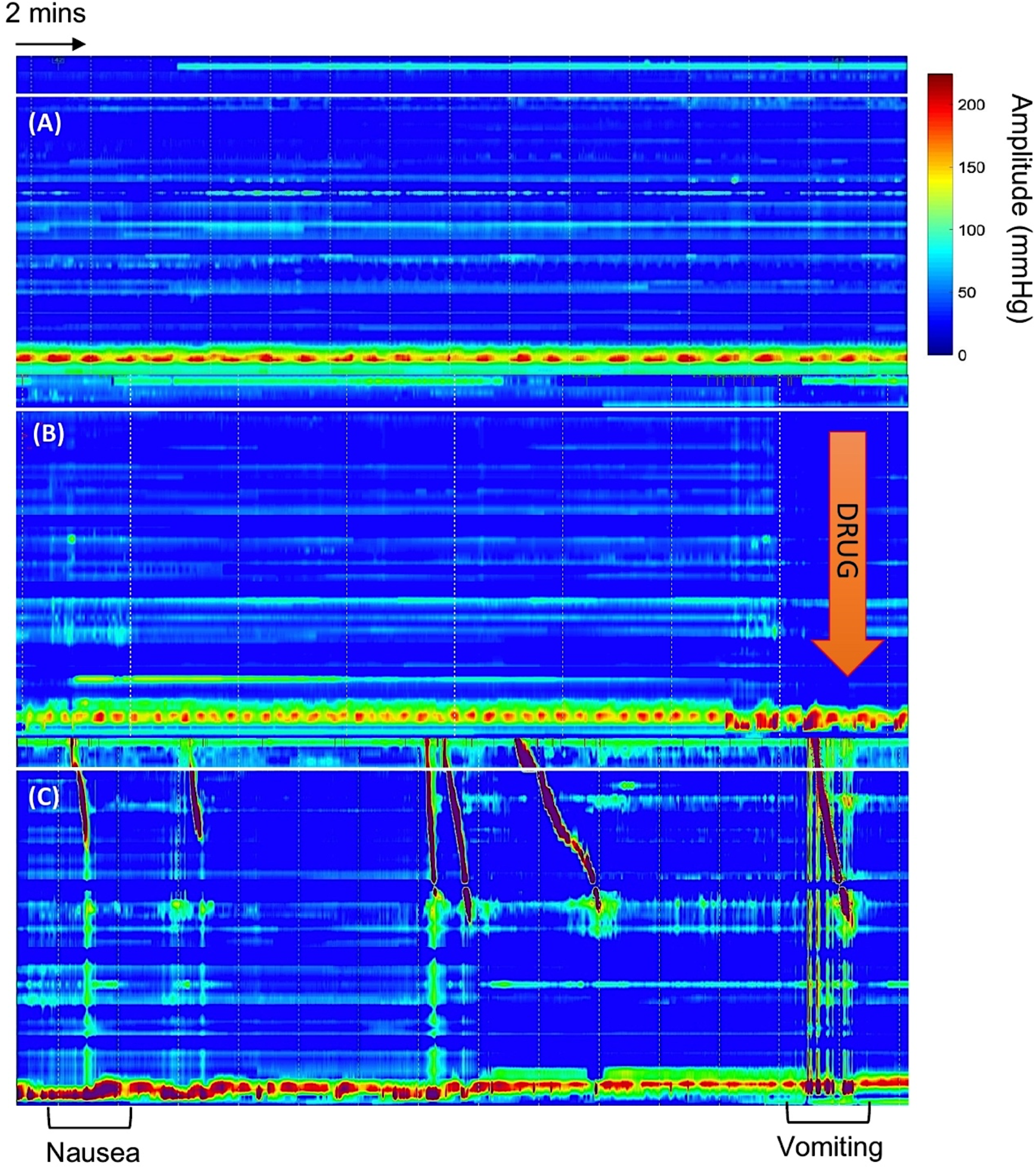
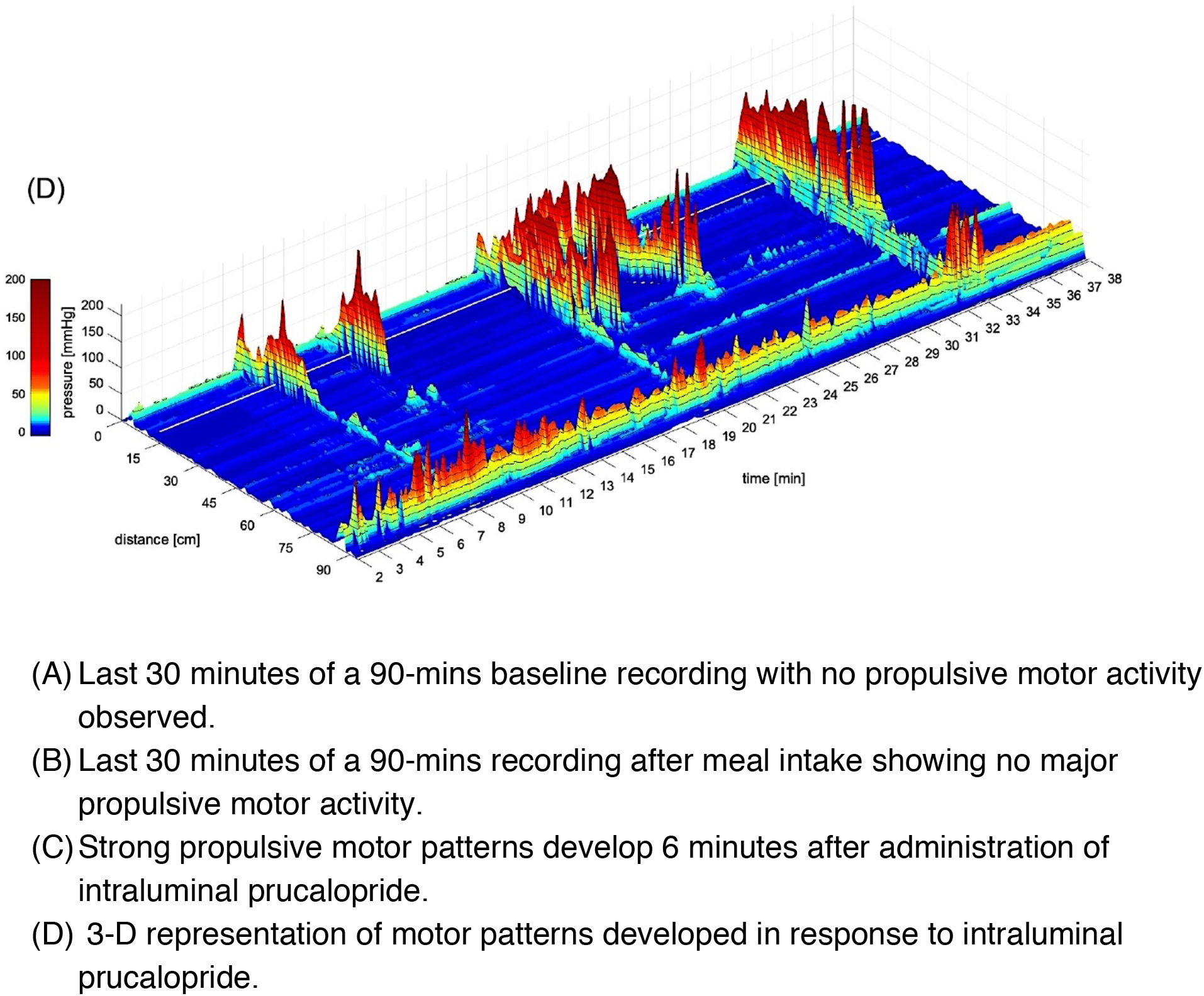
Intraluminal prucalopride resulted in significant propulsive motor activity compared with both baseline and meal response

**Table 1.** Characteristics of Motor Patterns in Response to Intraluminal Prucalopride

|  | HAPW | SPW | HAPW in HAPW-SPW | SPW in HAPW-SPW |
| --- | --- | --- | --- | --- |
| **Baseline (90 mins)** |  |  |  |  |
| **Frequency (cpm)** | ---<br>(n=0) | ---<br>(n=1) | ---<br>(n=0) | ---<br>(n=0) |
| **Mean Amplitude (mmHg)** | NA | 8.7 | NA | NA |
| **Duration (s)** | NA | 3.7 | NA | NA |
| **Velocity (cm s<sup>-1</sup>)** | NA | NA | NA | NA |
| **Meal (90 mins)** |  |  |  |  |
| **Frequency(cpm)** | ---<br>(n=0) | 0.03<br>(n= 3) | ---<br>(n=0) | ---<br>(n=0) |
| **Mean Amplitude (mmHg)** | NA | 7.1 ± 0.8<br>6.6 – 8 | NA | NA |
| **Duration (s)** | NA | 12.2 ± 2.5<br>10 – 15 | NA | NA |
| **Velocity (cm s<sup>-1</sup>)** | NA | NA | NA | NA |
| **Intraluminal Prucalopride 2 mg (60 mins)** |  |  |  |  |
| **Frequency(cpm)** | 0.07 <sup>*/#</sup><br>(n= 4) | 0.07 <sup>*</sup><br>(n=5) | 0.03 <sup>*/#</sup><br>(n= 2) | 0.03<sup>*/#</sup><br>(n= 2) |
| **Mean Amplitude (mmHg)** | 87.3 ± 6.4<br>80.9-93.7 | 13 ± 6.4 <sup>*/#</sup><br>7.3 – 24 | 69.9 ± 10.1<br>59.8-80 | 18.4 ± 9.8<br>11.4 – 25 |
| **Duration (s)** | 61.5 ± 37<br>23 – 112 | 6.2 ± 2<br>2.1 – 13.7 | 31 ± 4.4<br>28 – 34 | 6 ± 2<br>4.6 – 7.5 |
| **Velocity (cm s<sup>-1</sup>)** | 0.5 ± 0.4<br>0.2 – 1.15 | NA | 0.5 ± 0.01<br>0.50 – 0.53 | NA |

Values are mean ± SEM. Significance was determined using one-way ANOVA with multiple comparisons, \*compared to baseline (\* P< 0.05), # compared to meal (# P <0.05).

### Effects of Intraluminal Prucalopride on Propulsive Motor Patterns

Administration of intraluminal prucalopride in the proximal colon introduced HAPWs and HAPW-SPWs whereas none were present during the preceding 180 min of baseline and meal response (Figures 1C,D). Moreover, intraluminal prucalopride resulted in a significant increase of the SPW amplitude compared to both baseline and meal response (Table 1, Figure 2A). These effects were observed only 6 mins after administration of intraluminal prucalopride.

**Figure 2.**
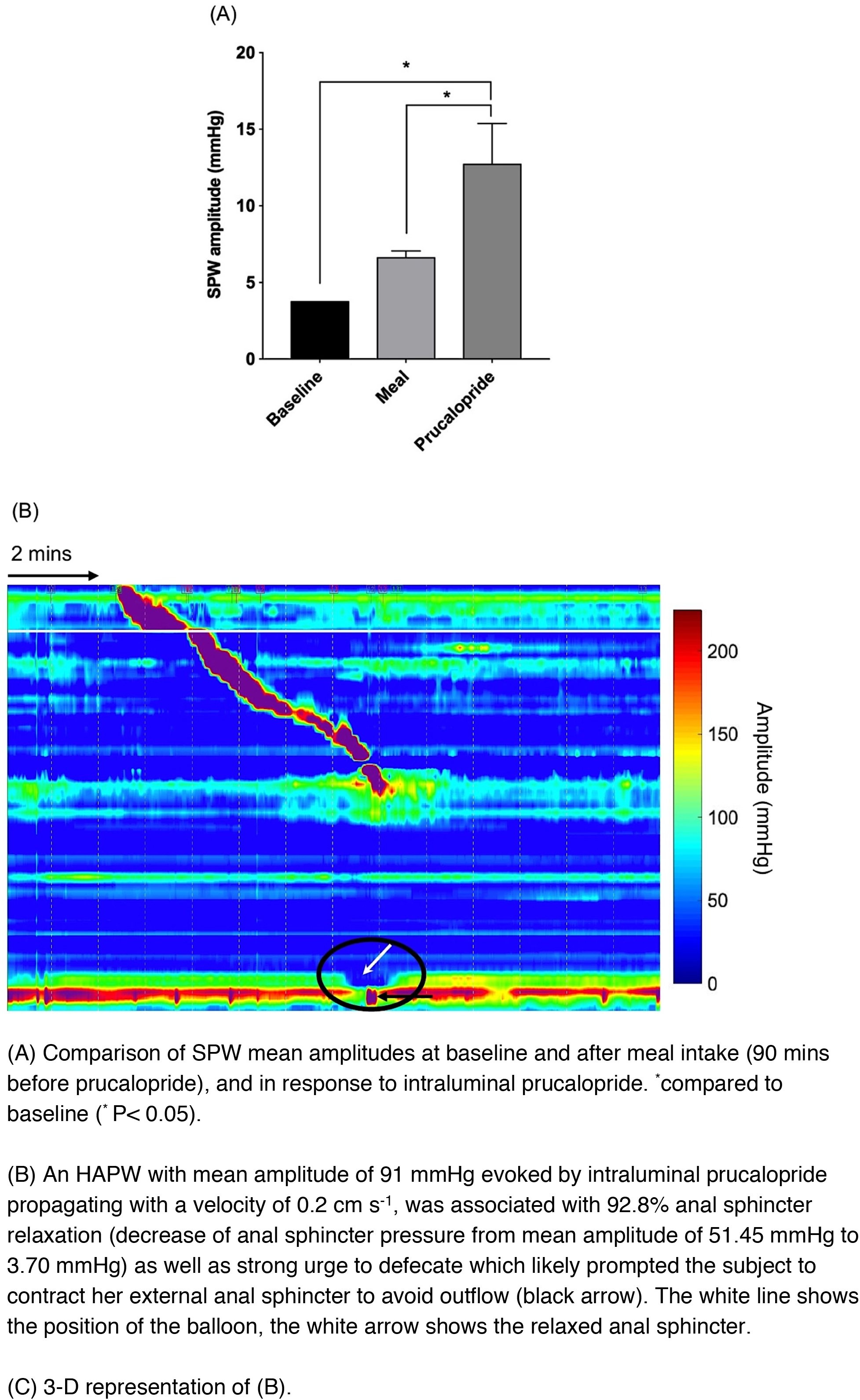
Motor patterns evoked by intraluminal prucalopride associated with significant anal sphincter relaxation, urge to defecate and increase in the amplitude of SPWs.

During the 30 min period after prucalopride administration, six HAPWs were generated, all belonging to category 2, that is, they started in the ascending colon and propagated to the transverse or descending colon with or without a change into an SPW. We have previously classified HAPWs based on their origin and place of termination in three distinct categories as described in Milkova et al. (37). The average amplitude of the HAPWs was 81.5 mmHg and the average HAPW Index was 2200 mmHg.m.s. The HAPW Index assesses the vigor with which a contraction occurs, and it was calculated as the multiplication of average amplitude within the 20-mmHg isobar, the length of the HAPW and the duration, expressed as mmHg.m.s. (37). The HAPWs here propagated with an average velocity of 0.53 ± 0.4 cm s^−1^. One HAPW was associated with urge to defecate (Figures 2B,C & 3). Two HAPWs without SPWs, were associated with significant, 78%, anal sphincter relaxation, lasting 26.5 ± 7 s.

**Figure 3.**
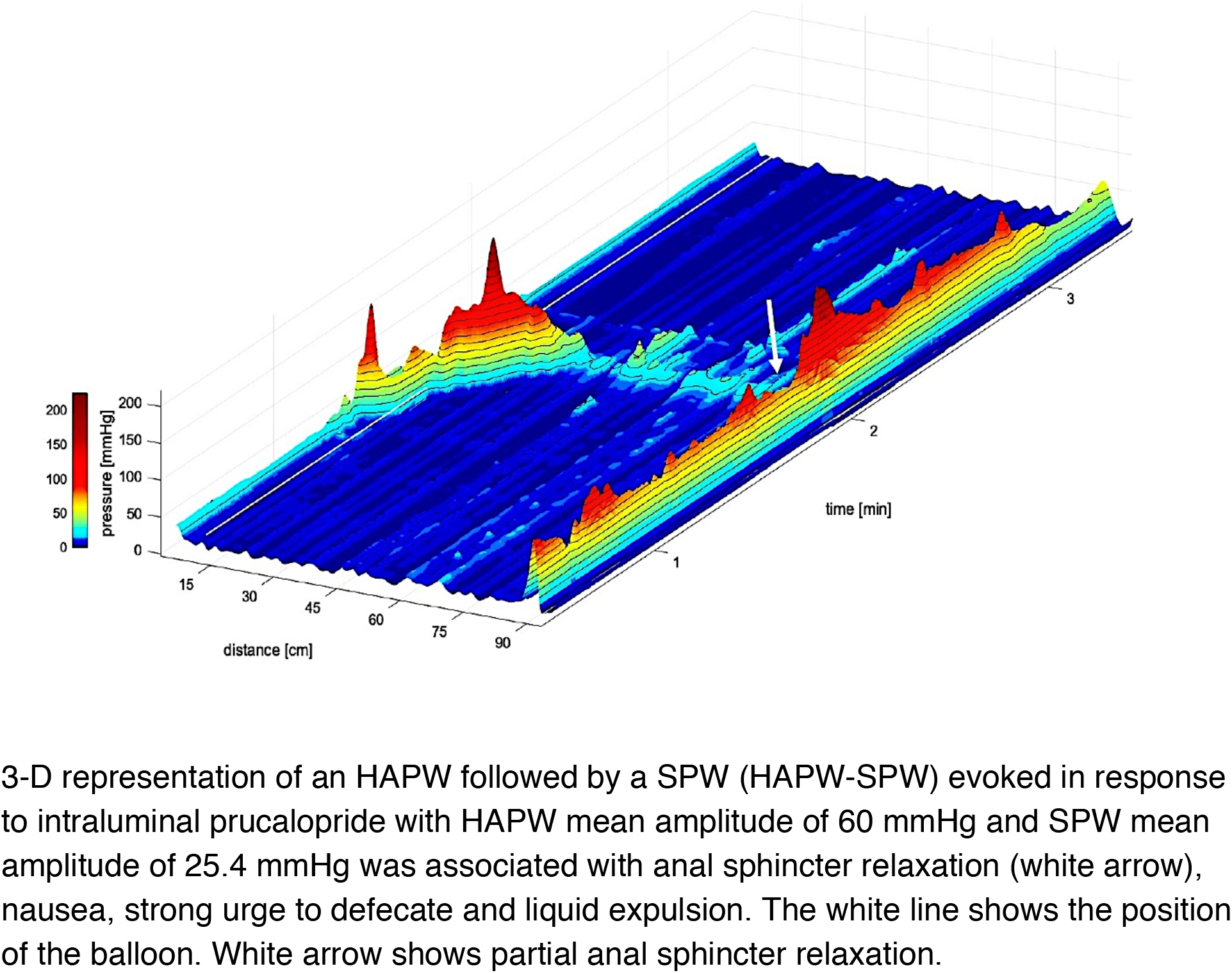
HAPW-SPWs evoked by intraluminal prucalopride could be associated with strong urge to defecate, anal sphincter relaxation and liquid expulsion.

Among the six HAPWs, two were HAPW-SPWs with the SPWs entering the rectum with an average amplitude of 18.4 ± 9.8 mmHg (Table 2), they were associated with liquid expulsion, urge to defecate and nausea (Figures 1, 2B, 3). These motor patterns were associated with 48% anal sphincter relaxation, from 63 to 33 mmHg.

**Table 2.**
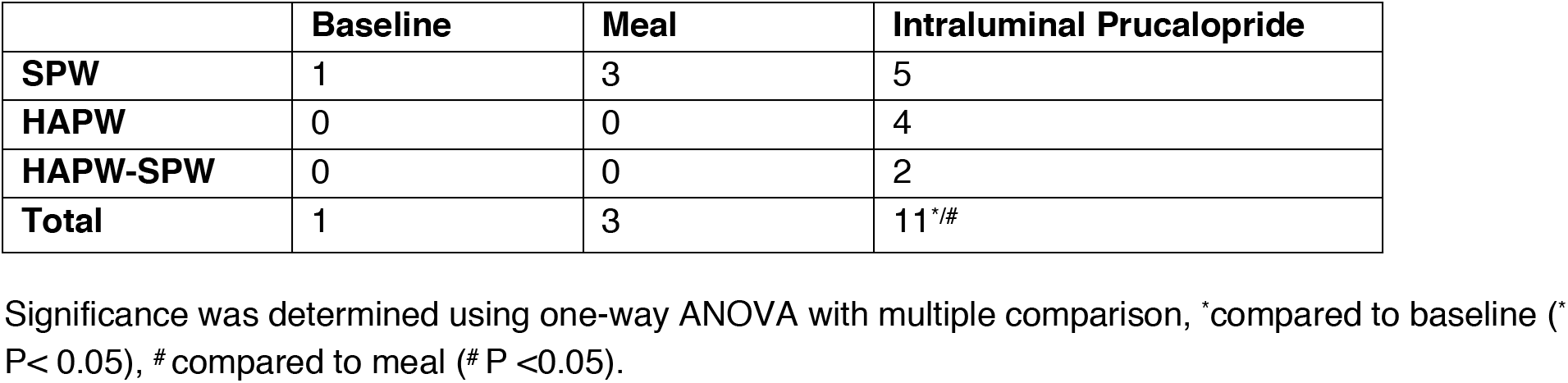
Total Occurrence of Propulsive Motor Patterns

**Table 3.** Anal Sphincter Relaxation Associated with Motor Patterns

|  | **Motor pattern mean amplitude (mmHg)** | **Anal sphincter mean amplitude before relaxation (mmHg)** | **Anal sphincter mean amplitude during relaxation (mmHg)** | **% of relaxation** | **Duration of relaxation (s)** | **Time of occurrence** |
| --- | --- | --- | --- | --- | --- | --- |
| **HAPW** |  |  |  |  |  |  |
| **Intraluminal prucalopride (n=2)** | 90.0 ± 1.1<br>88.9-91.1 | 40.5 ± 11<br>7.7 – 107 | 8.8 ± 5.1<br>3.6 – 44.3 | 78 | 26.5 ± 7 | When HAPWs entered the left colon |
| **SPW** |  |  |  |  |  |  |
| **Intraluminal prucalopride (n=4)** | 10 ± 1.10<br>7.30 – 12.40 | 31.60<br>15.40 – 56 | 13.10<br>3.60 – 45 | 60 | 13.3 | At the onset of SPW-cluster |
| **Baseline (n=1)** | 8.70<br>3.70 – 24 | 27.70<br>14 – 46 | 11.30<br>5 – 23 | 69 | 10.8 | At the onset of SPW |
| **Meal (n=3)** | 7.15 ± 0.80<br>6.60 – 8 | 26 ± 4.40<br>5.30 – 57.30 | 7.70 ± 8<br>2 – 20.30 | 87 | 15.6 ± 3.3 | At the onset of SPW |
| **HAPW-SPW (HAPW data)** |  |  |  |  |  |  |
| **Intraluminal prucalopride (n=2)** | 69.9 ± 10.1<br>59.8-80 | 62.50 ± 1<br>6 – 184 | 32.61± 12<br>6 – 101 | 48 | 25.8 ± 7.5 | At the onset of SPW |
| **HAPW-SPW (SPW data)** |  |  |  |  |  |  |
| **Intraluminal prucalopride (n=2)** | 18.4 ± 9.8<br>11.5 – 25.4 | 62.5 ± 1<br>6 – 184 | 32.6 ± 12<br>6 – 101 | 48 | 25.8 ± 7.5 | At the onset of SPW |

Values are mean ± SEM.

Five pan- colonic SPWs were evoked with an average amplitude of 13.0 ± 6.4 mmHg ranging between 7.3 – 24.0 mmHg with an average duration of 6.2 ± 2 s (Table 1). Vomiting associated pressure transients occurred at the end of the recording (Figure 1).

A SPW-cluster consisting of 4 pan-colonic SPWs was associated with significant anal sphincter relaxation. Anal sphincter did not recover in between the four SPWs. Before relaxation, the mean amplitude of the sphincter was 31.6 mmHg with maximum and minimum amplitudes of 56.0 and 15.4 mmHg respectively. During the relaxation phase, the anal sphincter amplitude decreased to 13.1 mmHg with maximum pressure of 45 and minimum pressure of 3.6 mmHg resulting in 60% relaxation (Table 2). One pan colonic SPW overlapped with an HAPW; therefore, anal sphincter relaxation could not be reliably attributed to either of the motor patterns.

The degree of relaxation associated with SPWs during baseline, meal and intraluminal prucalopride did not show any significant difference.

### Location of 5-HT and 5-HT_4_R Immunoreactivities in Colonic Epithelial Cells

To identify enterochromaffin cells, immunostaining with two types of 5-HT markers (polyclonal rabbit anti 5-HT and monoclonal mouse anti 5-HT) was performed which showed 100% colocalization in all sections of the colon. Immunostaining of monoclonal mouse anti 5-HT with both enzyme-induced epitope retrieval and heat-induced epitope retrieval showed similar results (P = 0.965). This verified the reliability of the mouse 5-HT antibody. 5-HT and 5-HT_4_R-positive cells were completely co-localized in the colonic epithelium in all sections. They were both scattered amongst other epithelial cells in transverse (Figures 4A,B), descending (Figure 4C), sigmoid colons (Figure 4D) and rectum (Figure 4E), mainly in the bottom crypts. A number of 5-HT-positive cells dispersed in the lamina propria were 5-HT_4_R negative (Figure 4B). These are likely mucosal 5-HT-positive mast cells.

**Figure 4.**
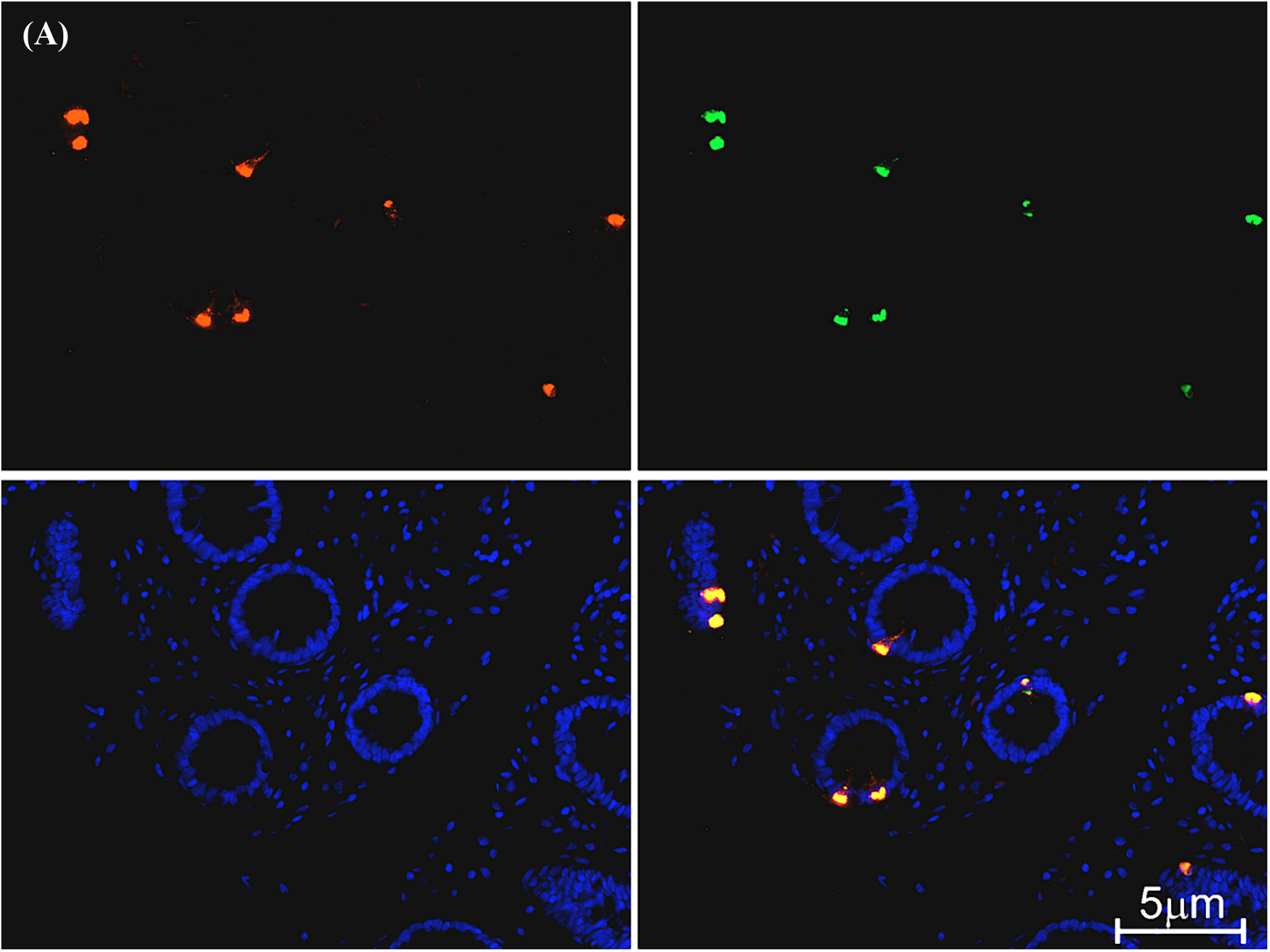
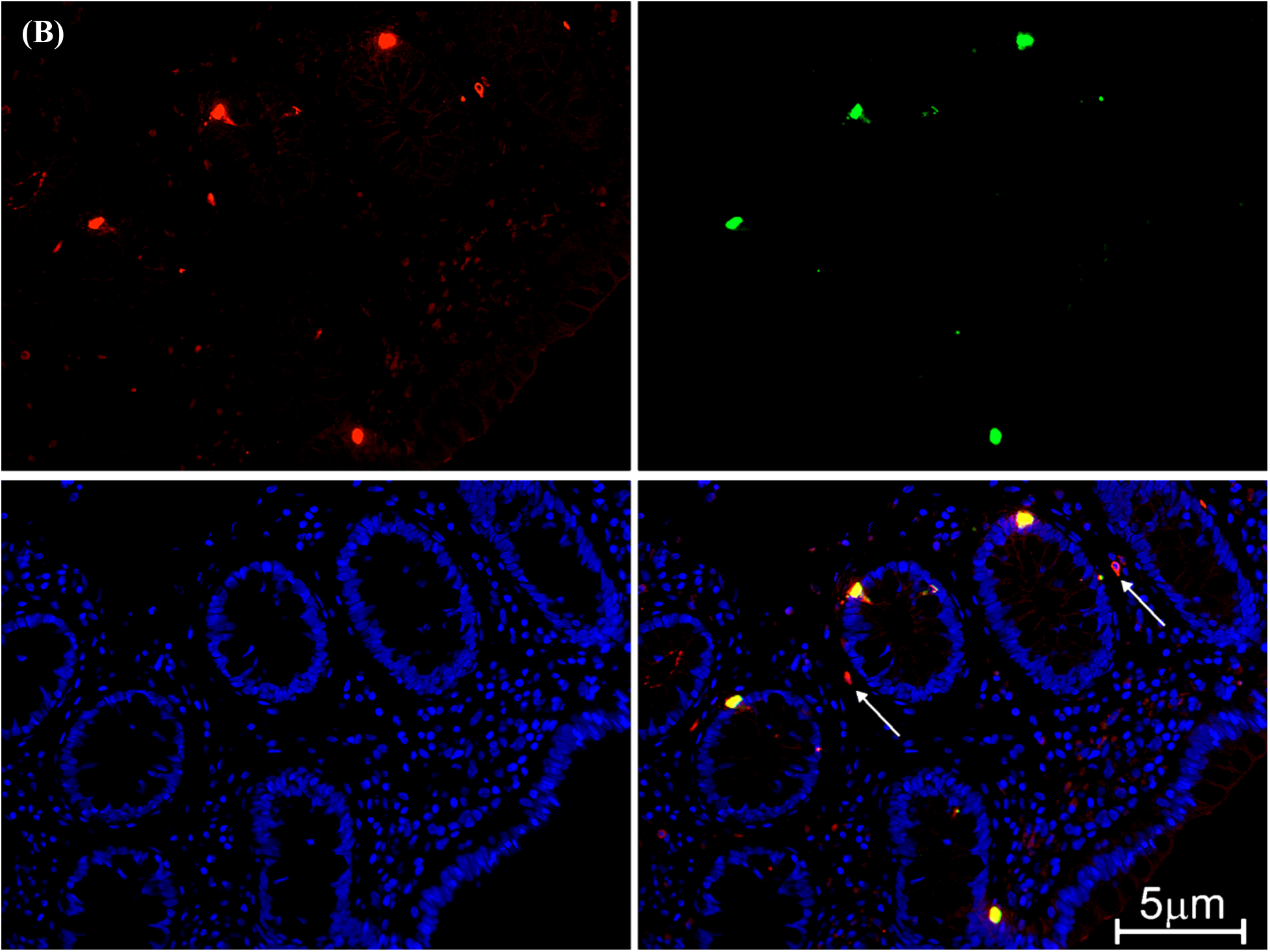
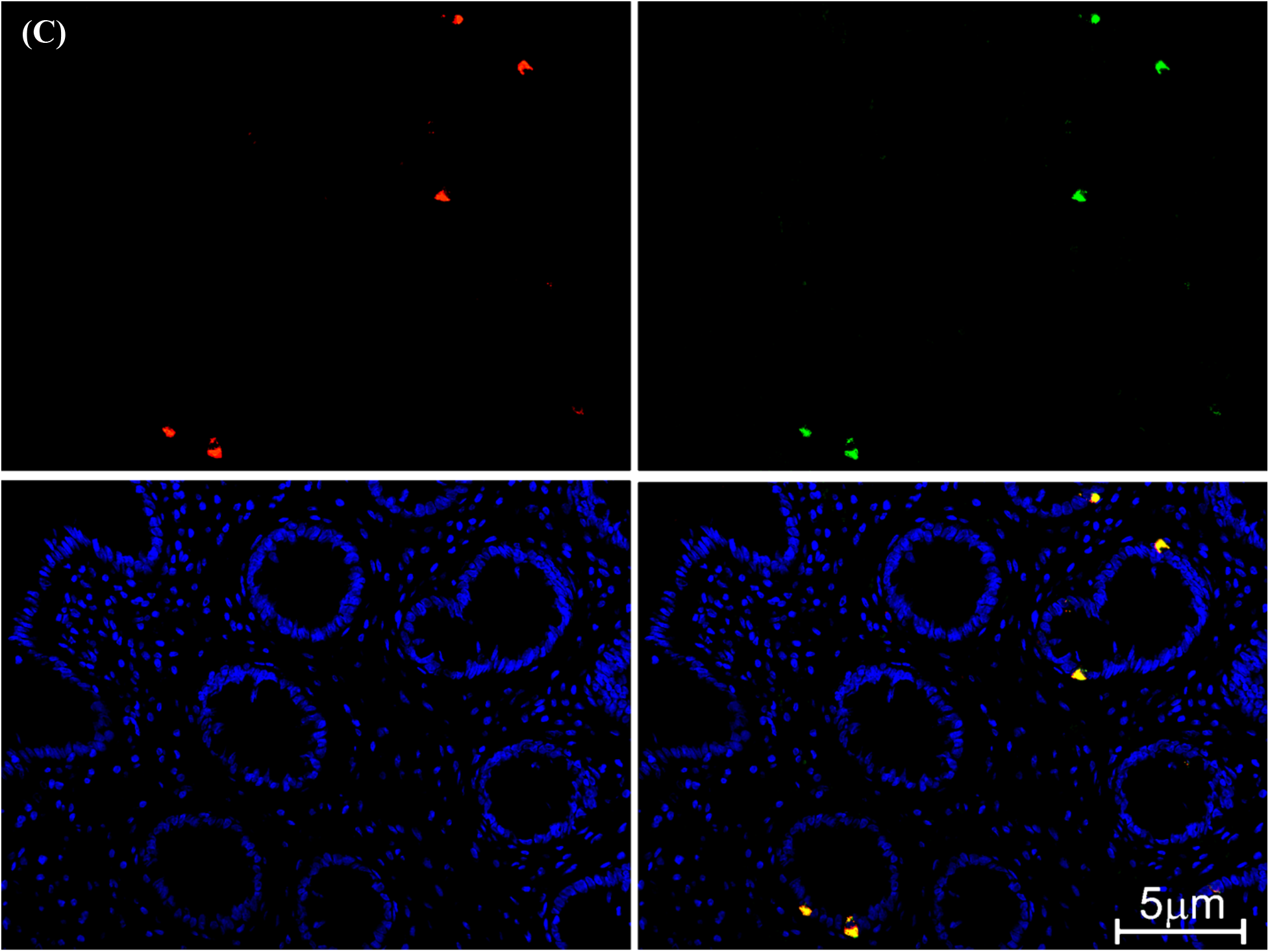
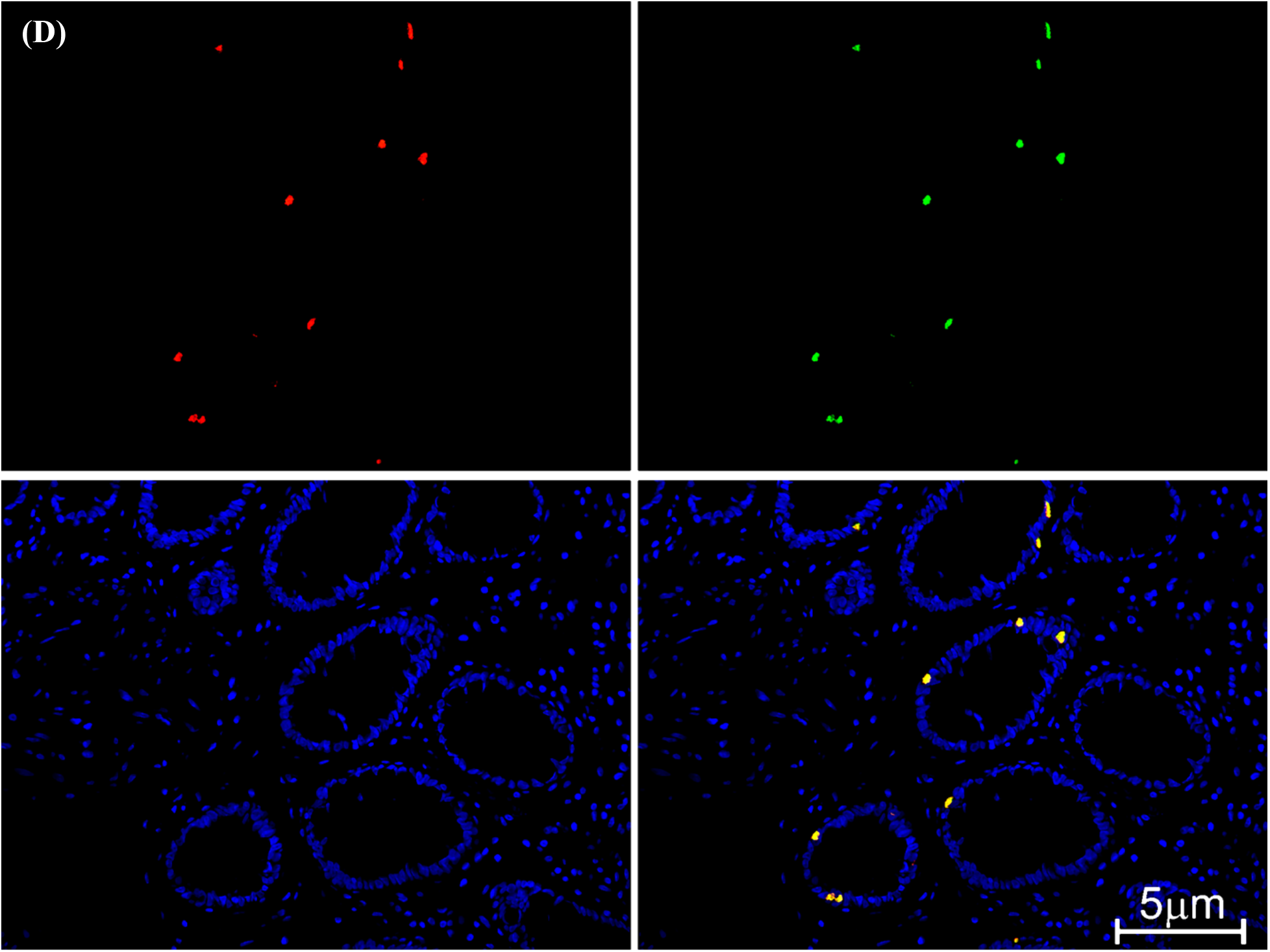
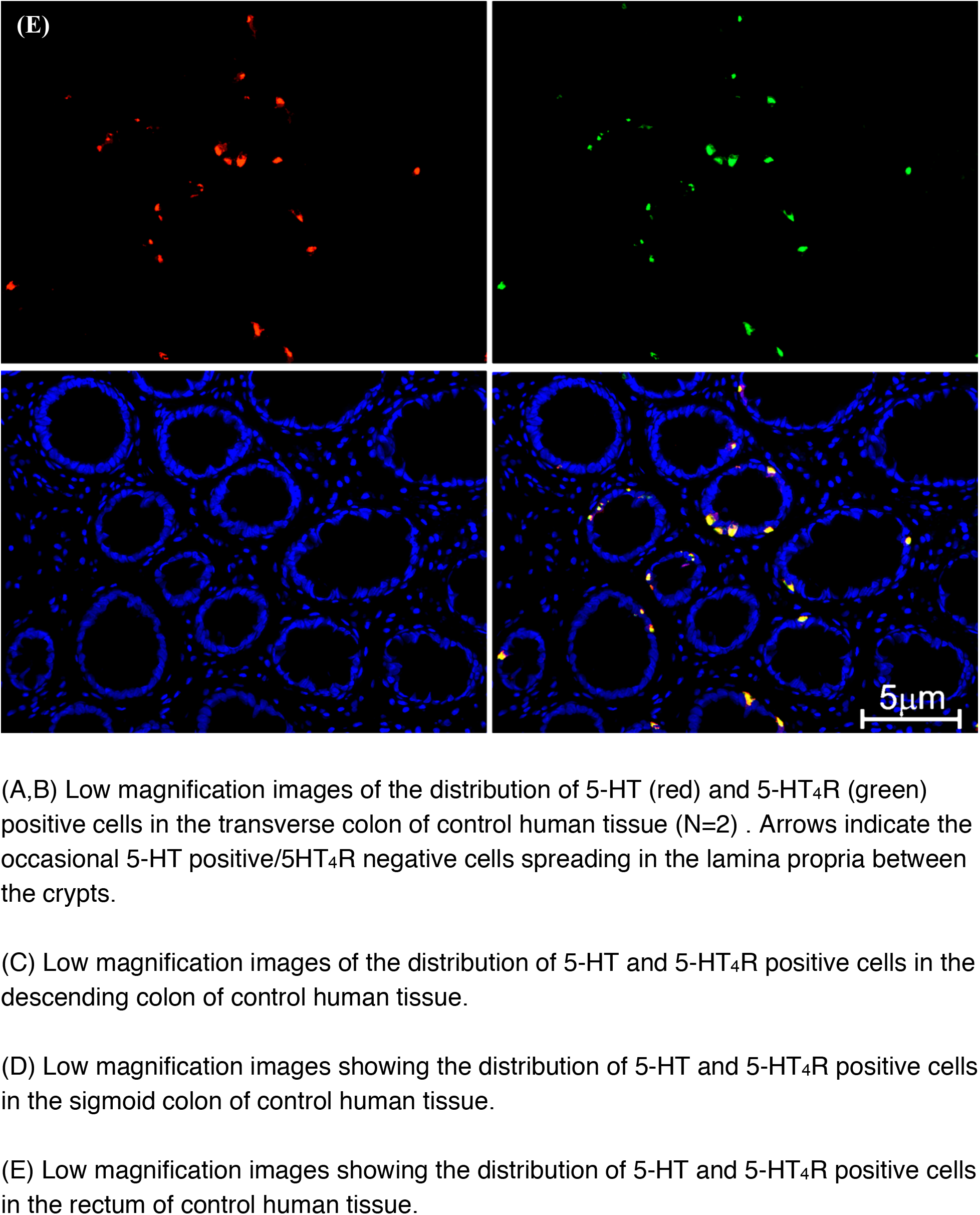
5-HT and 5-HT_4_R positive cells were completely co-localized in human colonic epithelium of all sections.

## DISCUSSION

### Baseline and Meal Activity

Baseline activity was low but not abnormal (37). In response to the meal, 3 SPWs occurred signifying a gastro-colonic reflex response which is a vagally-mediated awakening of the colon (1) (28) (37). In our study on HAPWs in healthy volunteers, 11% responded to a meal with only SPWs whereas 63% developed HAPWs, hence the response to the meal in this subject was low but not abnormal.

### Luminal Prucalopride

Luminal administration of prucalopride in the proximal human colon evoked powerful propulsive motor patterns and enhanced the amplitude of simultaneous pressure waves compared to both baseline and after meal. This response is similar to a typical response to luminal bisacodyl (37) (4). The induction of vomiting is a known effect of mucosal 5-HT receptor activation in the small bowel (48). The effect of prucalopride is likely mediated by 5-HT_4_Rs on enterochromaffin cells since we found an abundance of the 5-HT_4_R in the epithelial layer of biopsies of this patient and 100% colocalization of the receptor with 5-HT positive cells.

We have previously demonstrated that the major propulsive motor pattern in the rabbit colon, the colonic motor complex, manifests itself in different motor pattern configurations depending on the level of excitation of the colon. In the rabbit, intraluminal prucalopride, in a dose dependent manner, evoked several motor patterns starting with fast propagating contractions (45) that result in simultaneous pressure waves (39) to the most forceful propulsive contraction, the Long Distance Contraction (LDC), that results in high-amplitude propagating pressure waves (39), similar to the HAPW reported here. The rabbit study and the present study suggest that SPWs and HAPWs are part of the repertoire of propulsive activities in the human colon and that the SPWs are generally evoked at lower levels of excitation.

The HAPW is an essential component of the process of defecation and patients with constipation tend to have fewer HAPWs (16) (4, 40) (23). Reduction in occurrence of HAPWs can be accompanied by a decreased frequency of urge to defecate (34). Consistently, some patients with diarrhea-predominant irritable bowel syndrome (IBS-D) have been observed to generate more HAPWs compared with healthy subjects (25). Data reported here, indicate that luminal prucalopride can act as an effective prokinetic. Its effect appears to be similar to that of oral prucalopride (13).

### Mechanism of Action of Intraluminal Prucalopride

It is likely that intraluminal prucalopride induces propulsive motor patterns in humans by a sequence of events that start with 5-HT release from enterochromaffin cells leading to activation of mucosal endings of IPANs which communicate with neurons in the myenteric plexus. IPANs and vagal and pelvic afferent endings come into close proximity to the mucosal epithelium (50) (36) (18) which exposes them to chemicals released from enterochromaffin cells. Subsequently, propagation of excitation along the colon occurs through myenteric neuronal pathways in concert with myenteric ICC to activate longitudinal and circular muscles.

It has been suggested that the 5-HT_4_R is ubiquitously expressed by all epithelial cells in the murine colon. This was based on photomicrograph data from colonic sections of a 5-HT_4_R(BAC)-eGFP transgenic mouse in which cells express the 5-HT_4_R green fluorescent signal (36). In contrast, our data using immunohistochemistry revealed 100% co-expression of 5-HT and 5HT_4_R in human colonic epithelium throughout the colon, suggesting that enterochromaffin cells are the only epithelial cells expressing 5HT_4_Rs in the human colon. It is therefore likely that luminal prucalopride acts on these 5-HT_4_Rs.

### The Colo-Anal Reflex

The propagating motor patterns in the present study induced by prucalopride show a close temporal association with anal sphincter relaxation indicative of the colo-anal reflex, an independent neurally-controlled reflex involving autonomic sacral neural pathways (29, 30) (46) (43) (35) (37). The relaxation was often complete unlike the recto-anal inhibitory reflex (RAIR) which is characterized by a transient involuntary relaxation of the internal anal sphincter in response to rectal balloon distension. The internal anal sphincter will be inhibited by direct activation of intrinsic inhibitory nerves (12) (30) (42) and via parasympathetic activation of myenteric inhibitory nerves (20). Relaxation of the external anal sphincter can be achieved through decrease in the discharge frequency of sacral motor neurons innervating the sphincter (20). The pudendal nerve fibers that reach the external anal sphincter have cell bodies in the Onuf’s nucleus and can be stimulated by parasympathetic nerves, likely via interneurons, from the sacral defecation center (44) (31) (7) resulting in external anal sphincter relaxation.

### Clinical Relevance

The present study supports incorporation of prucalopride in colon-specific drug delivery systems to be used as a prokinetic for treating hypomotility disorders. Newer drug delivery systems targeting the colon use polysaccharide coatings that can be degraded only by the microorganisms existing in the colon which results in increased delivery success. With the advancement of nanotechnology, various forms of nanoparticle formulations for colon delivery have also been investigated (53) which could replace other types of colon-specific delivery systems in the future. This can be especially beneficial for high-risk populations such as the elderly and children.

## Data Availability

All data are in the manuscript

## Acknowledgements

This study was funded by the Canadian Institute of Health Research (PJT 152942) to Jan D Huizinga and an Ontario Graduate Scholarship to Mitra Shokrollahi. Ji-Hong Chen and Xuan-Yu Wang received salary support from the Farncombe Family Digestive Health Research Institute.

## Contributions of authors

JHC & JDH designed the study. JHC performed the HRCM. MS analyzed the HRCM data, made figures and tables and wrote the first draft of the manuscript. XYW performed the immunohistochemistry experiments. NM was involved in data comparison with other healthy volunteers. All authors contributed to data interpretation, edited the manuscript and approved the final version.

**Abbreviations**
5-HT: 5-hydroxytryptamine (serotonin)
HRCM: High-resolution colonic manometry
LDC: Long distance contraction
FPC: fast propagating contraction
HAPW: High-amplitude propagating pressure wave
SPW: simultaneous pressure wave.

